# Beyond posterior putamen lesions in post-stroke spasticity: widespread structural and functional breakdown across brain networks

**DOI:** 10.1101/2025.11.17.25340382

**Authors:** Antonio Jimenez-Marin, Gil De Sousa, Iñigo Tellaetxe-Elorriaga, Iñaki Escudero, Alberto Cabrera-Zubizarreta, Marimar Freijo, Pedro I Tejada, Ibai Diez, Asier Erramuzpe, Jesus M. Cortes

## Abstract

**Background:** Post-stroke spasticity (PSS) is a common and disabling motor complication whose neuroanatomical underpinnings remain incompletely understood. Beyond focal lesion localization, PSS likely arises from large-scale network disruptions involving both cortical and subcortical systems. Here, we combined functional and structural lesion network mapping (LNM) to identify the brain networks and white-matter tracts whose dysconnection best explains the presence and severity of PSS.

**Methods:** We analyzed 281 patients with hemorrhagic stroke, including 88 with PSS and 193 without PSS (NPSS). After matching for age, sex, and lesion size (N=81 PSS; N=60 NPSS), we computed functional LNM using normative connectomes from 1000 healthy participants of the Human Connectome Project and structural LNM using the HCP1065 tractography template. Group-level voxel-wise and tract-wise analyses identified regions and tracts with significant dysconnectivity differences. Finally, a ridge regression model with leave-one-out cross-validation assessed the relationship between dysconnectivity (both functional and structural) and spasticity severity, quantified by the Modified Ashworth Scale (MAS).

**Results:** Lesion-symptom mapping revealed that PSS was primarily associated with lesions in the posterior putamen. Structural LNM showed significantly higher tract dysconnectivity in PSS, particularly within the corticospinal, corticopontine, corticobulbar, corticostriatal, medial lemniscus, and superior thalamic radiation pathways (all p<0.001). Functional LNM analyses demonstrated widespread cortical dysconnectivity, with two complementary patterns: a Positive Dysconnected Network (PSS>NPSS) and a Negative Dysconnected Network (PSS<NPSS). Despite these bidirectional effects, overall dysconnection was consistently higher in the PSS group. The most affected networks included the cerebellum, sensorimotor, auditory, dorsal attention, default mode, and medial visual resting-state networks. Ridge regression analysis confirmed a strong association between MAS scores and the degree of dysconnectivity across these networks (R²=0.70).

**Conclusions:** Post-stroke spasticity is not the result of damage to a single locus but rather reflects the dysconnection of a distributed structural and functional network encompassing, not only motor, but also attentional and high order cognitive networks. These findings provide a network-based framework for understanding spasticity and support the use of lesion network mapping to predict and potentially guide targeted neuromodulation in stroke rehabilitation.

## Introduction

Spasticity is classically categorized within the upper motor neuron syndrome and is defined as a velocity-dependent increase in tonic stretch reflexes, reflecting hyperexcitability of spinal reflex pathways^1^ arising primarily from a loss of supraspinal inhibitory control over segmental motor circuits, rather than from changes in the peripheral musculature itself.

Post-stroke spasticity (PSS) affects 20% and 40% of stroke survivors and is one of the most disabling symptoms affecting patients’ functional independence in daily activities, thereby exerting a significant impact on the patient, the caregiver, and the healthcare system^2,3^. Early post-stroke management of spasticity is increasingly recognized as critical for optimizing recovery. Spasticity often emerges within days to weeks after stroke, and prompt intervention can attenuate its severity and forestall downstream complications. Indeed, early identification and treatment of high-risk patients has been linked to reduced long-term hypertonia and fewer secondary problems such as joint contractures, pain, and falls, which otherwise exacerbate disability^4^.

Traditionally, despite the high heterogeneity of lesions leading to PSS, clinical interpretation has tended to associate PSS with specific brain regions, resulting in a predominantly focal view of lesion studies. At the functional level, the most frequently reported affected region is the posterior putamen^5,6^, whereas at the structural level, spasticity has been primarily linked to damage in the corticospinal tract^7^.

However, most of the neurological symptoms arise from dysfunction within distributed brain networks connecting multiple regions. Indeed, Lesion Network Mapping (LNM) has shown in recent years that different lesions producing the same symptom are part of the same network^8^. LNM has explained brain injuries causing motor, non-motor, cognitive, and behavioral symptoms following stroke^9–15^. Within studies using LNM, networks have been found to be associated with symptoms such as hallucinations^16^, delusions^17^, abnormal movements^18,19^, pain^11,20^, coma^21^, and cognitive or social dysfunction^22,23^. More specifically, the LNM approach uses normative functional connectivity data from healthy populations to identify the brain networks that are typically connected to the patient’s lesion. Then, it is assumed that the lesion induces a “dysconnection” or alteration towards those networks. As a result, when analyzing a group of patients with anatomically heterogeneous lesions but sharing a common symptom, the convergence of dysconnected networks reveals the network substrate underlying that symptom. This paradigm represents a conceptual shift from traditional lesion localization and functional imaging toward a network-based understanding of brain function. The objective of the study is to determine the functional dysconnection of brain networks that underlie spasticity post-stroke.

## Methods

### Participants

This study was approved by the Clinical Research Ethics Committee of the Cruces University Hospital (code E19/52, PI: Jesus M Cortes). This is a retrospective study involving patients with stroke for assessing the relationship between brain imaging and different rehabilitation outcomes.

Eligible participants included all patients admitted to the Gorliz Hospital during the chronic-phase of stroke who underwent a rehabilitation program. A total of 281 hemorrhagic stroke patients were included in the study, with a mean age of 70.7 years (σ = 13.2, range 19 – 97), including 166 males. Of these, spasticity was present in 88 patients and absent in 193. Both groups were matched to be comparable in age, sex distribution, and lesion volume. After the matching process, final groups were N=81 patients with post-stroke spasticity (PSS) and N=60 patients with no PSS (NPSS). Modified Ashworth Scale (MAS)^24^ was available in 65 PSS patients. All the patients had a computed tomography (CT) acquired at the acute-phase hospital admission.

### Lesion segmentation and co-registration

Lesions were automatically segmented by finding the hyperdensities in the CT images. After that and following the recommendation of two experienced neuroradiologists (IE, AC), the lesion masks which were not well-defined were manually edited using 3D Slicer software (version 5.6.2; https://www.slicer.org/). To register the masks to the MNI152 template, we developed a tool (https://github.com/compneurobilbao/CTLesion2MNI152) based on^25,26^. Very briefly, our tool detects the skull on the CT based on the Hounsfield Units (HU > 100) and computes an affine registration matrix to the MNI152 skull. Then the tool extracts the brain (HU > 0, HU < 100) and applies the affine registration, with the addition of a nonlinear warp calculated without consideration of the lesion area for avoiding lesion-derived distortions that can worsen the registration quality.

### Structural lesion network mapping

To characterize the amount of lesion-induced dysconnectivity to white matter (WM) tracts, we used the HPC1065 tract atlas^27^. For each projection, association or cerebellar tract in the atlas, we used the MRtrix3 software package (v. 3.0_RC3) to exclude the fibers inside every lesion mask. Finally, we computed the ratio between the remaining fibers after exclusion and the total number of fibers for every lesion/tract in the atlas. Due the atlas being lateralized, we averaged the dysconnectivity values of each tract across the left and right hemispheres, yielding a single value per tract. This approach reduces data dimensionality and simplifies the problem of multiple comparison corrections.

### Functional lesion network mapping

To successfully map stroke lesions to their respective dysconnectivity patterns, resting-state functional MRI (rs-fMRI) data from healthy controls (*n* = 1000) were obtained from the Human Connectome Project (HCP) dataset^28^. The data was preprocessed with ICA-FIX^29^, and additionally we applied global signal regression, band-pass filtering between 0.01-0.08Hz and spatial smoothing with FWHM of 6mm. The functional connectivity maps for each stroke patient were generated through seed-based connectivity (SBC) analysis following the pipeline described in Jimenez-Marin et al. (2022). Summarizing, the lesion mask of each stroke patient registered to the MNI152 template was used as the seed region for SBC analyses, which were conducted separately for each HCP subject. Pearson correlation coefficients (r) were computed between the mean timeseries of the seed region (obtained by averaging voxel-wise time-series within each lesion) and the time-series of all other brain voxels. To improve Gaussianity in the data, these r-values were Fisher-transformed using the inverse hyperbolic tangent function. This process resulted in a 3D brain map of z-values for each stroke lesion, generated independently for each HCP subject. A one-sample t-test was then applied across the 1000 individual connectivity maps to derive a final functional dysconnection map for each lesion, yielding a group-level statistical representation of lesion-induced functional dysconnectivity. The pipeline is openly available at https://github.com/compneurobilbao/lnm.

### Statistical Analysis

#### Demographics and behavioral differences between groups

Group differences (PSS vs. NPSS) in age, lesion volume and structural dysconnectivities were assessed using Wilcoxon rank-sum test for two samples. Sex differences were assessed using the χ2 test of independence of variables.

#### Lesion symptom mapping

Lesion locations were compared between groups. To do that, we assumed that the occurrence of spasticity is hemisphere-independent, so we flip the right-hemisphere lesions to the left for having less comparisons and increase statistical power. Then we fit a general linear model (GLM) for each voxel in the left hemisphere comparing both groups. Finally, we included as significant every voxel with (false discovery rate) FDR p-value < 0.01.

#### Functional Lesion network mapping

Lesion-induced functional dysconnectivity maps were compared between SP and NSP groups. We fit a GLM for each voxel in the gray matter comparing both groups. We considered as significant every voxel with FDR p-value < 0.001. Additionally, to reduce the number of possible false positive voxels, we applied the Monte Carlo simulation cluster-wise correction implemented in the AFNI v23.0.04 software package. We ran 10,000 iterations to estimate the probability of false positive clusters with a p value < 0.05. The maps of group differences were then described by calculating the overlap between the resting state networks (RSN) and also with brain maps of 22 neuro-behavioural meta-analytic concepts. The RSN maps were derived following (Tahedl & Schwarzbach, 2023), adding the dorsal attention, salience and language networks using the masks included in CONN toolbox (https://web.conn-toolbox.org/). We then compared the median dysconnectivity inside the RSN masks between groups. Bonferroni correction was finally applied to correct for multiple comparisons.

#### Lesion network maps associations with spasticity severity

In the PSS group, we fit a Ridge linear model with the spasticity severity (MAS) scale as the independent variable. To predict MAS, the selected features were the median dysconnectivities of RSNs and the tract dysconnectivities (only the ones showing significant differences for RSNs and tracts). The effect of lesion size was also regressed out from the independent variable and features, given its statistical dependence and the fact that larger lesions inherently produce greater dysconnection. We also applied leave-one-out cross-validation to test the model, reducing the overfitting. Finally, SHAP values^31^ were reported to evaluate the feature importance ranking.

## Results

Two different approaches based on LNM were applied to investigate the neural network basis of PSS. The first approach, functional LNM, was used to analyze differences in the dysconnection of functional brain networks between the PSS and NPSS groups. The second approach, structural LNM, focused instead on the dysconnection of major white matter tracts between groups. The methodological framework followed in this study is illustrated in Figure 1.

**Figure 1.**
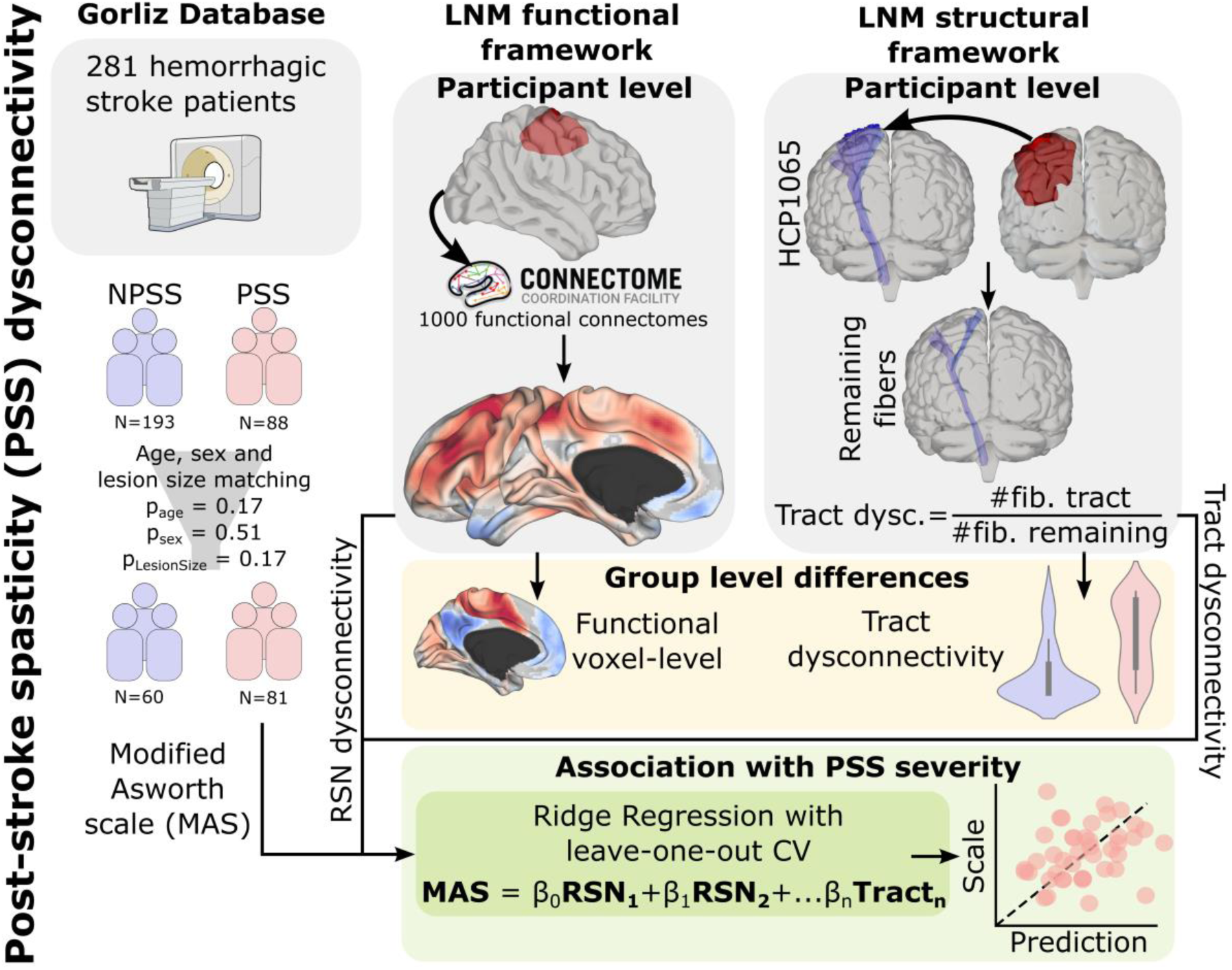
Methodological pipeline for lesion network mapping (LNM), providing network structural and functional dysconnectivity, and its association with post-stroke spasticity (PSS) severity. The study included 281 patients with hemorrhagic stroke recruited in the Neurorehabilitation Gorliz Hospital (Bizkaia, Spain). Among them, 193 patients did not present PSS (NPSS) and 88 did (PSS). After age, sex, and lesion size matching, 60 NPSS and 81 PSS patients were used for our analyses. Functional LNM was computed at the participant level using normative functional connectivity data from 1000 healthy subjects provided by the HCP Connectome Coordination Facility. Structural LNM was computed using the HCP1065 tractography template, quantifying tract dysconnectivity as the ratio between the number of affected fibers and the number of remaining fibers. At the group level, statistical comparisons were performed on voxel-wise functional dysconnectivity maps and on tract-wise dysconnectivity distributions to identify networks and tracts showing significant between-group differences. Finally, to assess the association between dysconnectivity and PSS severity (measured by the Modified Ashworth Scale, MAS), a ridge regression model with leave-one-out cross-validation was fitted, including as predictors the median dysconnectivity values of resting-state networks (RSNs) and tracts that showed significant group differences.

### Participant Characteristics

Both stroke groups (PSS and NPSS) were matched for age, sex, and lesion volume. The mean age was 64.2 ± 11.2 years in the PSS group and 66.5 ± 10.7 years in the NPSS group (p = 0.17). Males represented 72% (n = 58) of the PSS group and 65% (n = 39) of the NPSS group (p = 0.51). Mean lesion volumes were 15.1 ± 17.4 cm³ and 13.5 ± 19.1 cm³ in the PSS and NPSS groups, respectively (p = 0.17). The modal score on the Modified Ashworth Scale (MAS) in the PSS group was 2. Table 1 summarizes the demographic characteristics of both groups.

**Table 1:** Demographic and behavioural data.

| *Variable, units* | *SP* | *NSP* | *T* | *p* |
| --- | --- | --- | --- | --- |
| *Age, years* | 64.2 (11.2) | 66.5 (10.7) | -1.38 | 0.17 |
| *Males, n(%)* | 58 (72) | 39 (65) | 0.43 | 0.51 |
| *Lesion Volume (cm<sup>3</sup>)* | 15.1 (17.4) | 13.5 (19.1) | 1.38 | 0.17 |
| *MAS, n* | 2, 65 |  |  |  |

### Anatomical Differences in Lesion Location Between PSS and NPSS

Figure 2 shows the spatial brain maps of lesion location for both groups PSS and NPSS and the statistically significant differences between them, after multiple comparison corrections. Lesion location statistical differences, a.k.a Lesion Symptom Mapping (LSM), were found mainly in the posterior part of the putamen, in agreement with previous studies^5^.

**Figure 2.**
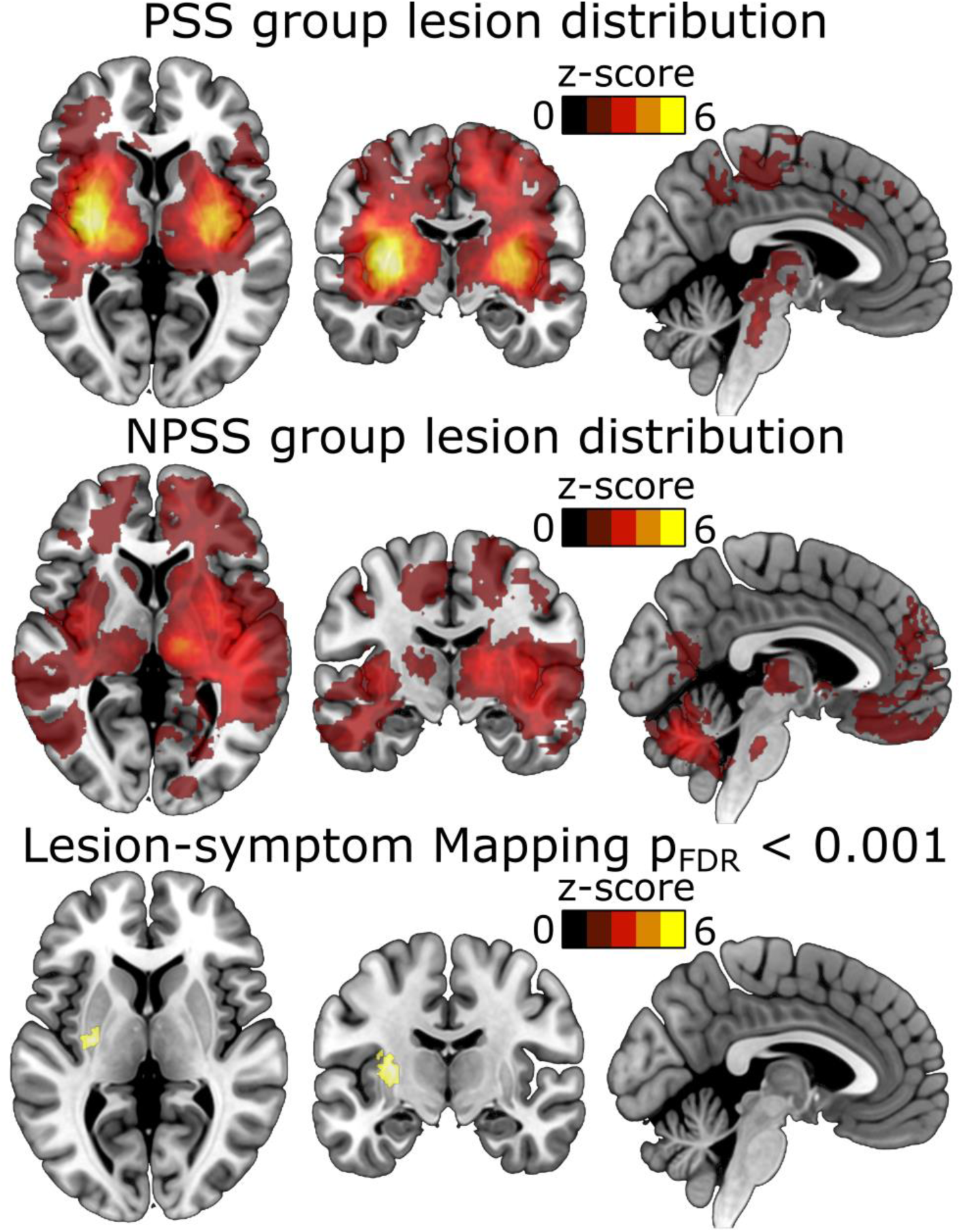
Lesion location distributions and lesion–symptom mapping results. Voxel-wise lesion overlap maps are shown for patients with post-stroke spasticity (PSS, top row) and those without spasticity (NPSS, middle row). In both groups, lesion recurrence is represented as z-score heatmaps. The NPSS group exhibited a more spatially heterogeneous lesion distribution, whereas the PSS group showed higher lesion overlap in subcortical and periventricular regions. The bottom row shows the results of the voxel-based lesion–symptom mapping analysis identifying regions significantly associated with PSS (p-FDR < 0.001). The most significant cluster was located in the posterior putamen, indicating that damage in this region is strongly associated with the presence of post-stroke spasticity.

### Distinct White Matter Dysconnectivity Patterns Underlying PSS and NPSS

Group differences in the amount of tract dysconnectivity were found in 9 tracts out of 39, after Bonferroni corrections. In all cases, higher dysconnectivity values were found in the PSS group as compared to NPSS (Figure 3). Significant differences were observed across several projection tracts, reported here in descending order of dysconnection strength (T value): corticospinal tract (T = 6.45, p < 0.001), superior corticostriatal tract (T = 6.22, p < 0.001), parietal corticopontine tract (T = 5.07, p < 0.001), frontal corticopontine tract (T = 4.64, p < 0.001), corticobulbar tract (T = 4.49, p < 0.001), posterior corticostriatal tract (T = 3.96, p = 0.003), medial lemniscus (T = 3.45, p = 0.022), and superior thalamic radiation (T = 3.32, p = 0.036). Furthermore, the association tract extreme capsule also showed increased dysconnectivity (T = 4.41, p < 0.001).

**Figure 3.**
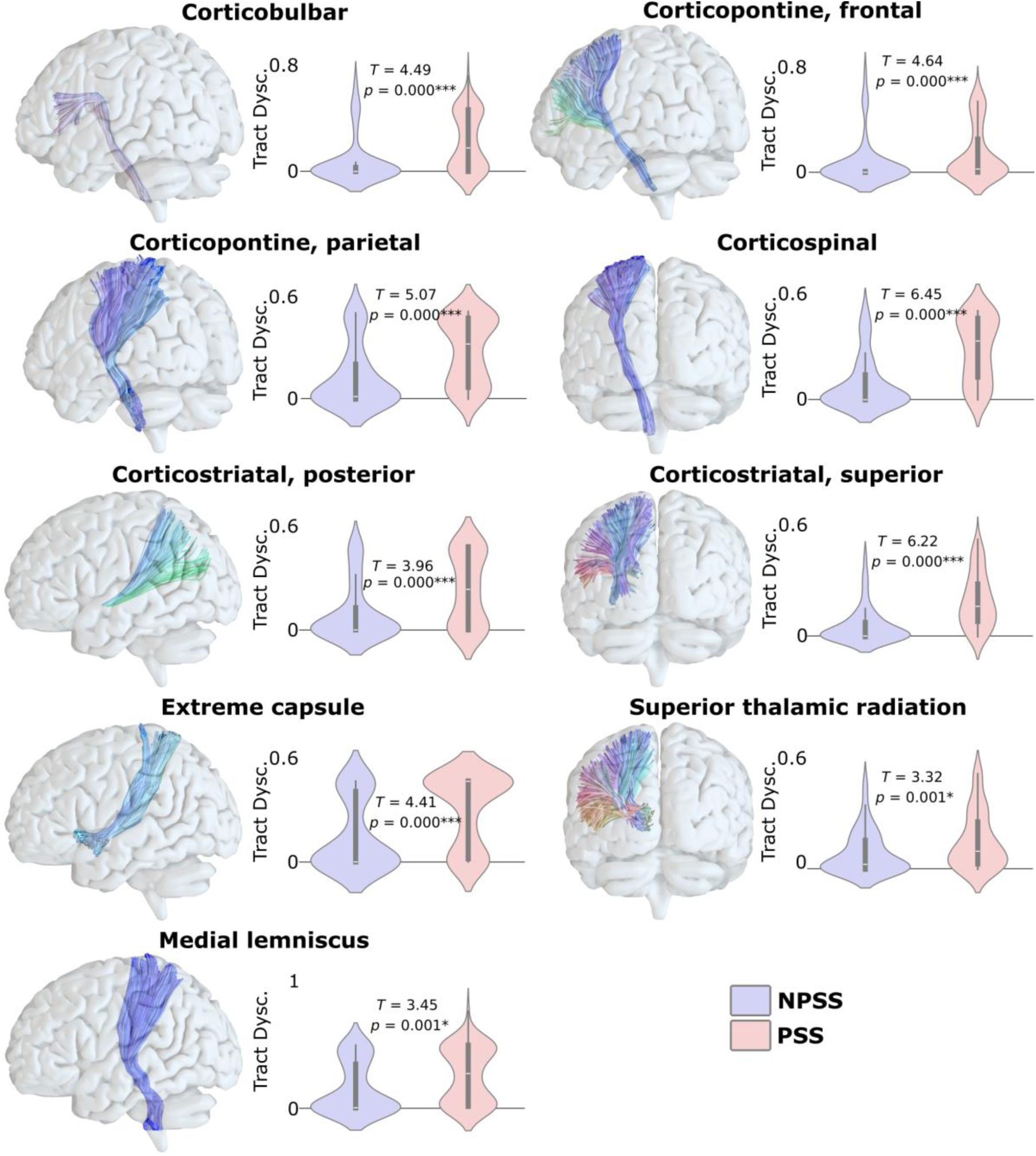
Group differences in tract-level structural dysconnectivity. Only white-matter tracts showing significant group differences after Bonferroni correction are displayed. Each panel depicts the corresponding tract within the MNI template brain and the distribution of tract dysconnectivity values across patients with post-stroke spasticity (PSS) and without spasticity (NPSS). Tract colors follow the conventional DTI orientation scheme: blue indicates superior–inferior, red left–right, and green anterior–posterior directions. Statistical comparisons were conducted using two-sample *t*-tests, reporting *T* and uncorrected *p*-values. Significance after Bonferroni correction is denoted as follows: n.s. (*p* > 0.05); * (*p* < 0.05); ** (*p* < 0.01); *** (*p* < 0.001). The tracts showing greater dysconnectivity in the PSS group included the corticospinal, corticopontine (frontal and parietal parts), corticobulbar, corticostriatal (posterior and superior parts), medial lemniscus, extreme capsule, and superior thalamic radiation, consistent with damage affecting both projection and association pathways.

### Distinct Functional Network Disruptions Underlying PSS and NPSS

PSS patients showed increased functional dysconnectivity compared with NPSS. This was observed for both positive contrasts (PSS > NPSS) and negative contrasts (PSS < NPSS). These two-sided differences corresponded to two distinct networks. The positive dysconnected network (PDN), characterized by higher dysconnectivity in the PSS group (PSS > NPSS), primarily involved motor and attention networks. In contrast, the negative dysconnected network (NDN), showing greater negative dysconnectivity in the PSS group (PSS < NPSS), was mainly associated with the default mode network (DMN) (Figure 4).

**Figure 4.**
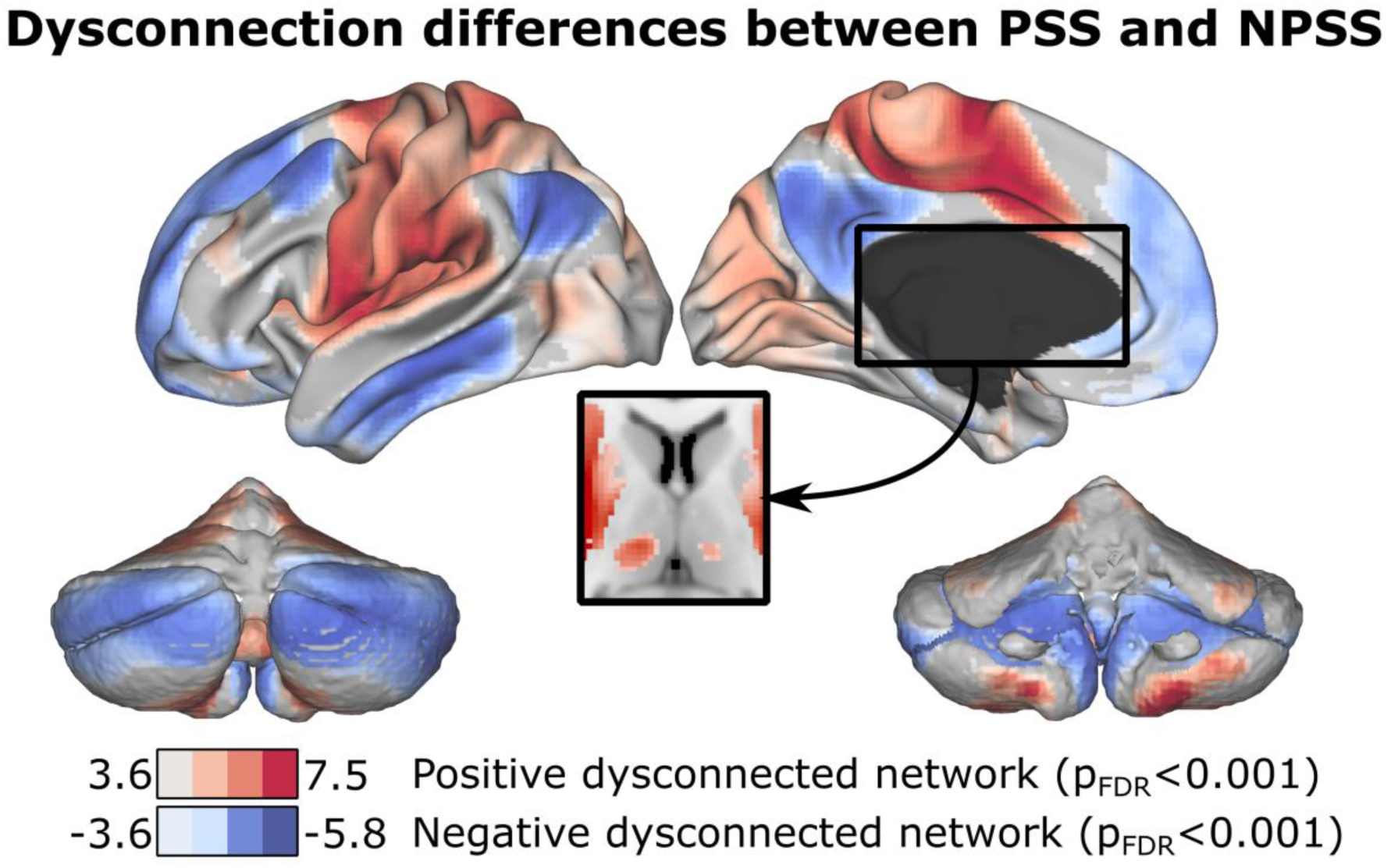
Functional dysconnectivity differences between PSS and NPSS groups. Voxel-wise comparisons of lesion network mapping (LNM) maps revealed widespread gray matter dysconnectivity differences between patients with post-stroke spasticity (PSS) and those without spasticity (NPSS), thresholded at *p*-FDR < 0.001. Two main spatial patterns were identified: regions showing greater dysconnectivity in the PSS group (PSS > NPSS), displayed in red, defined as the *Positive Dysconnected Network* (PDN); and regions showing reduced dysconnectivity in the PSS group (PSS < NPSS), displayed in blue, defined as the *Negative Dysconnected Network* (NDN). Importantly, irrespective of the network type, the PSS group exhibited overall greater functional dysconnection compared to the NPSS group, indicating a global increase in network-level dysconnectivity associated with spasticity.

Specifically, when looking at the overlapping with classical RSNs, we found high overlaps and significant Bonferroni-corrected differences in the amount of dysconnectivity across several networks (Figure 5). The medial visual network showed a 56.8% overlap with the PDN (T = 3.24, p = 0.001), while the lateral visual network overlapped by 32.8% with the PDN (T = 2.89, p = 0.004). The default mode network (DMN) exhibited a 78.8% overlap with the NDN (T = -4.46, p < 0.001). The cerebellar network had a 48.6% overlap with the PDN (T = 3.33, p = 0.001), and the sensorimotor network showed a nearly complete overlap of 99.1% with the PDN (T = 4.23, p < 0.001). The auditory network overlapped by 71.6% with the PDN (T = 4.34, p < 0.001), whereas the right frontoparietal network had a 43.8% overlap with the NDN (T = -2.96, p = 0.003). Finally, the dorsal attention network showed an 87.6% overlap with the PDN (T = 4.59, p < 0.001).

**Figure 5.**
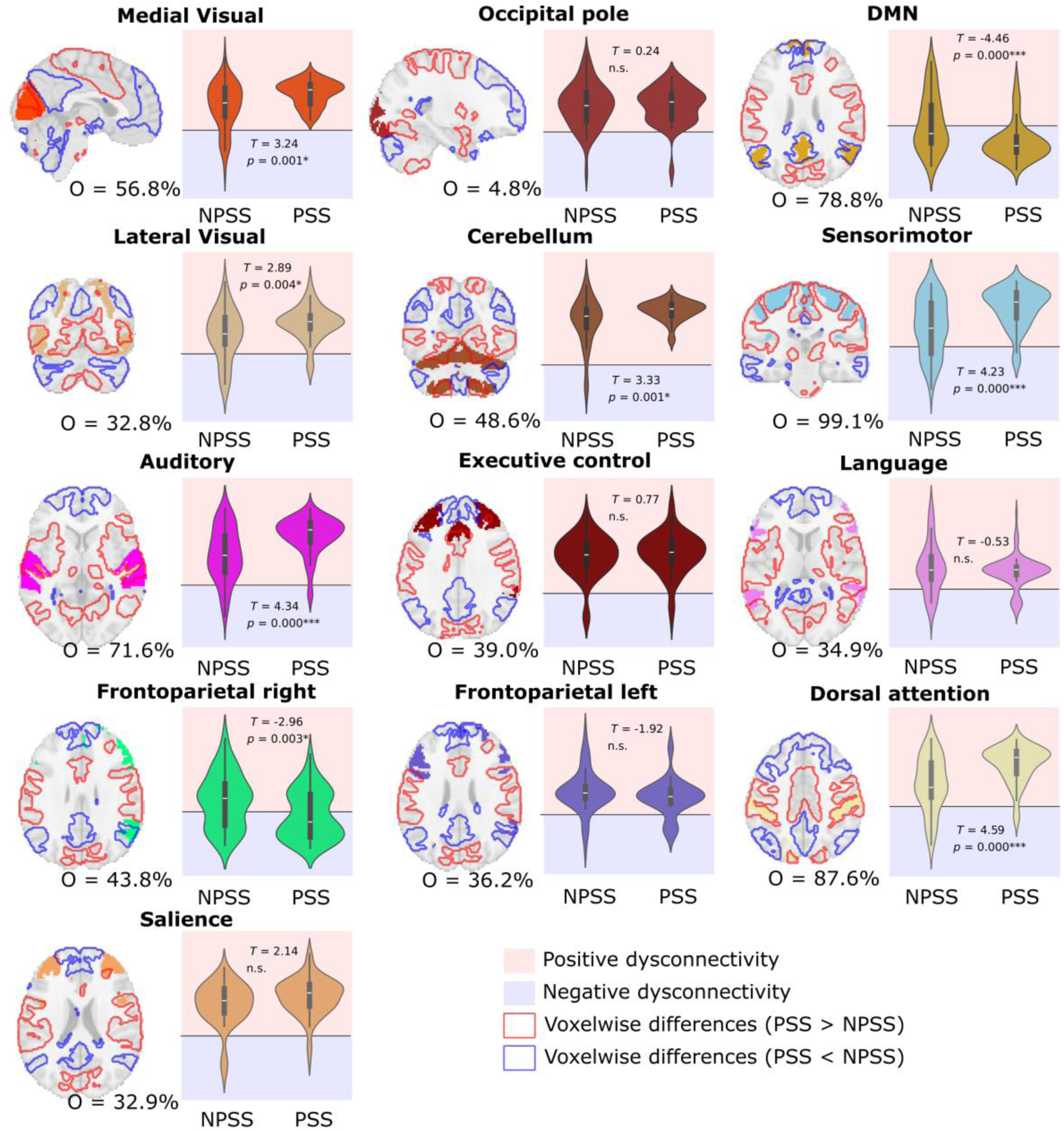
Overlap between dysconnectivity differences and resting-state networks (RSNs). The spatial overlap (O) between voxel-wise dysconnectivity differences and 13 canonical RSNs is shown, expressed as the percentage of RSN voxels overlapping with regions showing significant group differences. For each RSN, the solid-colored regions correspond to the network mask, while red and blue contours indicate areas with significantly higher (PSS > NPSS) or lower (PSS < NPSS) dysconnectivity, respectively. Violin plots display the distribution of median dysconnectivity values across participants within each RSN mask for both PSS and NPSS groups. Red and blue shaded areas correspond to positive and negative dysconnectivity, respectively. Statistical comparisons were performed using two-sample *t*-tests, reporting *T* and uncorrected *p*-values. Significance after Bonferroni correction is denoted as: n.s. (*p* > 0.05); * (*p* < 0.05); ** (*p* < 0.01); *** (*p* < 0.001). Networks showing significantly greater dysconnectivity in PSS included the cerebellum, sensorimotor, auditory, dorsal attention, default mode, medial and lateral visual, and right frontoparietal RSNs.

By using normative data to compare lesion connections between the PSS and NPSS groups, we have learned that the differences between groups extend across multiple functional networks. Moreover, in all these networks, the PSS group consistently showed a higher degree of dysconnectivity, affecting both positive and negative connections.

Alternatively, one might ask the following question: if we consider the region of interest revealed by the Lesion Symptom Mapping analysis as a seed (cf. Figure 2, bottom panel) and compute its connectivity to the whole brain in the same normative dataset, do we obtain results similar to those observed in the group differences from the LNM? That is, how much the LSM seed resulting from group-level differences in lesion-location associations resembles the group-level differences obtained from the LNM connectivity profile of each patient’s lesion with the rest of the brain in the normative dataset? Interestingly, we found that 69% of the variance in the LNM dysconnectivity differences could be explained by the LSM-based connectivity map. These results demonstrate that, although the connectivity pattern from the LSM seed partially resembles the LNM group difference maps, the two approaches still capture fundamentally distinct aspects of brain network disruption caused by brain injury.

### Association Between Network Dysconnectivity Patterns and MAS Severity

We fitted a Ridge regression model to predict the MAS score using the functional dysconnectivity of those RSNs that showed significant group differences in the LNM maps, together with the amount of tract dysconnectivity of significant white matter tracts as features. This resulted in a total of 17 features, achieving a highly significant performance with R² = 0.70. After performing leave-one-out cross-validation (LOO-CV) to reduce over-fitting, R² decreased to 0.47, while maintaining a linear relationship between the set of features and the MAS score. We also tested the LOO-CV model trained only with the functional features (R² = 0.42). When trained only with the structural features, we obtained R² = 0.27. Therefore, although structural dysconnectivity alone is a moderate predictor of spasticity severity, functional dysconnectivity is a much better predictor. Moreover, when both structural and functional features are combined, they jointly provide the best prediction of spasticity and its severity.

Furthermore, we computed SHAP values (Sv) to assess the relative importance of each feature in predicting MAS severity. Among the functional features, the cerebellar RSN was the most influential (Sv = 1.12), followed by the medial visual (Sv = 0.56), default mode (Sv = 0.53), right frontoparietal (Sv = 0.45), auditory (Sv = 0.38), dorsal attention (Sv = 0.33), lateral visual (Sv = 0.16), and sensorimotor (Sv = 0.15) networks. Regarding structural features, the most relevant tract was the medial lemniscus (Sv = 0.25), followed by the corticopontine tract (parietal part, Sv = 0.13) and the corticospinal tract (Sv = 0.09).

## Discussion

### Summary of findings

This study investigated the neuroanatomical and network correlates of post-stroke spasticity by combining voxel-based LSM, functional and structural LNM in a large cohort of hemorrhagic stroke survivors, where we identified the posterior putamen as the main lesion site associated with spasticity. Beyond lesion location, PSS was characterized by widespread functional and structural dysconnectivity affecting several structural pathways and well-known resting state networks. Structural analyses revealed significant dysconnectivity differences in nine white-matter tracts, while functional analyses revealed two large-scale dysconnectivity patterns: A Positive Dysconnected Network (PDN, PSS > NPSS) and a Negative Dysconnected Network (NDN, PSS < NPSS). Overall, PSS patients showed consistently higher network dysconnection, which predicted spasticity severity with high accuracy (R² = 0.70).

### Interpretation

Our findings indicate that PSS emerges from distributed supraspinal network dysfunction rather than isolated spinal or focal cortical damage. The LSM results confirmed the posterior putamen and its connections as the primary structures associated with spasticity, consistent with^5^. The putamen, a core nucleus of the basal ganglia, regulates motor initiation, inhibition, and learning through cortico-basal ganglia-thalamo-cortical loops^32^. Damage to this region may disrupt the balance between direct and indirect basal ganglia pathways^33^, producing abnormal excitatory-inhibitory motor control and, consequently, spasticity. Its additional role in sensorimotor integration and procedural automation^34^ further supports its involvement in tone regulation and reflex modulation.

Structurally, the tracts showing significant dysconnectivity—the corticospinal, corticostriatal, corticopontine (frontal and parietal), corticobulbar, medial lemniscus, superior thalamic radiation, and extreme capsule—represent a combination of projection and association fibers central to motor function. The corticospinal tract, essential for voluntary movement^35^, showed the largest statistical differences. The corticostriatal and corticopontine tracts link cortical motor areas with the basal ganglia and cerebellum, supporting fine coordination and motor learning^36,37^. The corticobulbar tract contributes to facial and cranial motor control^38^, while the medial lemniscus and superior thalamic radiation relay proprioceptive and sensory feedback crucial for movement regulation^35,39^. Although the extreme capsule is typically associated with language processing^40^, its dysconnection may reflect fronto-insular diaschisis or shared fiber crossings adjacent to periventricular lesions.

Functionally, dysconnectivity was widespread across gray matter, forming two complementary patterns. The PDN included areas with positive dysconnection, mainly encompassing the sensorimotor, dorsal attention, cerebellar, auditory, and visual networks. The NDN included regions with negative dysconnectivity, notably in the default mode and right frontoparietal networks. Despite these opposite directions, the global dysconnection load was consistently higher in PSS, reinforcing the idea of network-level vulnerability. The sensorimotor network, with nearly complete overlap (∼100%) with the PDN, highlights its dominant role in spasticity. The dorsal attention network’s disruption is consistent with its integrative sensory–motor function^41^. In contrast, DMN and frontoparietal changes may reflect compensatory or inhibitory network reorganization^42–44^. The involvement of visual and auditory networks may represent multimodal contributions to motor control and feedback, in line with findings of visually responsive neurons in the putamen^34^.

### Comparison with previous work

Our findings replicate and extend the results of the first lesion network study on PSS^6^, which identified a bilateral putamen–globus pallidus hub in patients with spasticity but not in those without. We confirm this hub’s involvement and expand its network context to include several functional and structural networks. Importantly, we demonstrate that approximately 70% of the variance in LNM results can be explained by the LSM-derived connectivity, suggesting that lesion dysconnectivity-symptom relationships are largely mediated through network patterns anchored in key subcortical structures.

Moreover, unlike prior studies limited to functional LNM, our approach combined structural and functional dysconnectivity and introduced a cross-validated predictive model linking these measures to clinical severity (MAS). This methodological innovation provides a quantitative framework for mapping connectome-level injury using only lesion masks, allowing us to perform advanced network analyses in clinical cohorts where fMRI or DWI data are not routinely available.

### Clinical implications

Conceptualizing PSS as a network-level disorder has direct implications for prognosis and treatment. Identifying key nodes such as the posterior putamen, sensorimotor cortex, and cerebellum may guide the development of targeted neuromodulation interventions, including transcranial magnetic or direct current stimulation. Moreover, early identification of patients at risk based on lesion-network profiles could optimize rehabilitation timing and intensity. From a practical point of view, LNM-based methods require only routinary clinical imaging, making them accessible to most hospital settings and scalable to large patient cohorts.

Rehabilitation strategies designed to enhance supraspinal connectivity—such as connectome-guided stimulation or circuit-based motor training—could help restore functional integration between motor, cerebellar, and attention networks, potentially reducing spasticity and improving motor outcomes.

### Limitations and future directions

Several limitations should be acknowledged. First, although our sample is (as far as we know) the largest PSS cohort studied with LNM, replication in independent and multicentric datasets might be useful for generalizability of our results. Second, because resting-state fMRI and diffusion MRI were unavailable in our clinical cohort, we relied on normative connectomes; patient-specific data would allow direct validation of inferred dysconnections. Third, this study focused on hemorrhagic stroke, and generalization to ischemic lesions remains to be tested. Finally, our cross-sectional design limits causal inference; longitudinal imaging and follow-up of spasticity evolution are needed to determine whether network alterations precede or follow clinical onset.

Future research should investigate whether modulation of the identified networks (particularly the cerebellum, DMN, and putamen) can effectively reduce spasticity and whether connectomic biomarkers can predict recovery trajectories.

## Conclusion

Post-stroke spasticity arises from the disruption of a distributed network involving the posterior putamen and its structural and functional connections with motor, sensory, attention, and cerebellar systems. This dysconnection model bridges lesion localization with large-scale network dysfunction, offering a mechanistic explanation for tone abnormalities after stroke. Recognizing spasticity as a network-based disorder provides a rationale for precision rehabilitation and targeted neuromodulation aimed at restoring supraspinal connectivity and improving functional recovery.

## Data Availability

All data is available upon reasonable request.

## Acknowledgements

JMC acknowledges financial support from the Spanish Ministry of Health (PI22/01118) and Basque Ministry of Health (2023111002 & 2022111031). JMC and AE are both funded by the Spanish Ministry of Science (grant PID2023-148012OB-I00). ID was supported by the Spanish Ministry of Science with the grant PID2023-150633OA-I00. AE and ID are both funded by the Spanish Ministry of Science and Innovation grants RYC2021-032390-I and RYC2022-035429-I, respectively. JMC, AE, and ID are funded by Ikerbasque: The Basque Foundation for Science. MMF acknowledges financial support from Carlos III Institute of Health (grant RICORS-Ictus RD24/0009/0004).

## References

1. Lance J. symposium synopsis, in felman RG, Young RR, Koella WP (eds): spasticity: Disordered Motor Control Chicago. *Year b Med Publ*. 1980;494.

2. Ganapathy V, Graham GD, DiBonaventura MD, Gillard PJ, Goren A, Zorowitz RD. Caregiver burden, productivity loss, and indirect costs associated with caring for patients with poststroke spasticity. Clin Interv Aging. 2015;10:1793–1802. doi:10.2147/CIA.S91123

3. Zorowitz RD, Gillard PJ, Brainin M. Poststroke spasticity: sequelae and burden on stroke survivors and caregivers. Neurology. 2013;80(3 Suppl 2):S45–52. doi:10.1212/WNL.0b013e3182764c86

4. Wissel J, Verrier M, Simpson DM, et al. Post-stroke spasticity: predictors of early development and considerations for therapeutic intervention. PM R. 2015;7(1):60–67. doi:10.1016/j.pmrj.2014.08.946

5. Lee KB, Hong BY, Kim JS, et al. Which brain lesions produce spasticity? An observational study on 45 stroke patients. Bergsland N, ed. PLOS ONE. 2019;14(1):e0210038. doi:10.1371/journal.pone.0210038

6. Qin Y, Qiu S, Liu X, et al. Lesions causing post-stroke spasticity localize to a common brain network. Front Aging Neurosci. 2022;14:1011812. doi:10.3389/fnagi.2022.1011812

7. Huang L, Yi L, Huang H, Zhan S, Chen R, Yue Z. Corticospinal tract: a new hope for the treatment of post-stroke spasticity. Acta Neurol Belg. 2024;124(1):25–36. doi:10.1007/s13760-023-02377-w

8. Fox MD. Mapping Symptoms to Brain Networks with the Human Connectome. N Engl J Med. 2018;379(23):2237–2245. doi:10.1056/NEJMra1706158

9. Aerts H, Fias W, Caeyenberghs K, Marinazzo D. Brain networks under attack: robustness properties and the impact of lesions. Brain. 2016;139(12):3063–3083. doi:10.1093/brain/aww194

10. Boes AD, Prasad S, Liu H, et al. Network localization of neurological symptoms from focal brain lesions. Brain J Neurol. 2015;138(Pt 10):3061–3075. doi:10.1093/brain/awv228

11. Burke MJ, Joutsa J, Cohen AL, et al. Mapping migraine to a common brain network. Brain. 2020;143(2):541–553. doi:10.1093/brain/awz405

12. Darby RR, Joutsa J, Fox MD. Network localization of heterogeneous neuroimaging findings. Brain. 2019;142(1):70–79. doi:10.1093/brain/awy292

13. Jimenez-Marin A, De Bruyn N, Gooijers J, et al. Multimodal and multidomain lesion network mapping enhances prediction of sensorimotor behavior in stroke patients. Sci Rep. 2022;12(1):22400. doi:10.1038/s41598-022-26945-x

14. Joutsa J, Corp DT, Fox MD. Lesion network mapping for symptom localization: recent developments and future directions. Curr Opin Neurol. 2022;35(4):453–459. doi:10.1097/WCO.0000000000001085

15. Salvalaggio A, De Filippo De Grazia M, Zorzi M, Thiebaut de Schotten M, Corbetta M. Post-stroke deficit prediction from lesion and indirect structural and functional disconnection. Brain. 2020;143(7):2173–2188. doi:10.1093/brain/awaa156

16. Kim NY, Hsu J, Talmasov D, et al. Lesions causing hallucinations localize to one common brain network. Mol Psychiatry. 2021;26(4):1299–1309. doi:10.1038/s41380-019-0565-3

17. Rhee J, Ferguson M, Bonkhoff A, et al. Localizing Delirium through Automated Lesion Segmentation and Neural Network Mapping (N1.001). Neurology. 2023;100(17_supplement_2):3220. doi:10.1212/WNL.0000000000203110

18. Fasano A, Laganiere SE, Lam S, Fox MD. Lesions causing freezing of gait localize to a cerebellar functional network. Ann Neurol. 2017;81(1):129–141. doi:10.1002/ana.24845

19. Joutsa J, Horn A, Hsu J, Fox MD. Localizing parkinsonism based on focal brain lesions. Brain. 2018;141(8):2445–2456. doi:10.1093/brain/awy161

20. Kim NY, Taylor JJ, Kim YW, et al. Network Effects of Brain Lesions Causing Central Poststroke Pain. Ann Neurol. 2022;92(5):834–845. doi:10.1002/ana.26468

21. Fischer DB, Boes AD, Demertzi A, et al. A human brain network derived from coma-causing brainstem lesions. Neurology. 2016;87(23):2427–2434. doi:10.1212/WNL.0000000000003404

22. Cotovio G, Talmasov D, Barahona-Corrêa JB, et al. Mapping mania symptoms based on focal brain damage. J Clin Invest. 2020;130(10):5209–5222. doi:10.1172/JCI136096

23. Darby RR, Horn A, Cushman F, Fox MD. Lesion network localization of criminal behavior. Proc Natl Acad Sci. 2018;115(3):601–606. doi:10.1073/pnas.1706587115

24. Bohannon RW, Smith MB. Interrater reliability of a modified Ashworth scale of muscle spasticity. Phys Ther. 1987;67(2):206–207. doi:10.1093/ptj/67.2.206

25. Kuijf HJ, Biesbroek JM, Viergever MA, Biessels GJ, Vincken KL. Registration of Brain CT Images to an MRI Template for the Purpose of Lesion-Symptom Mapping. In: Shen L, Liu T, Yap PT, Huang H, Shen D, Westin CF, eds. Multimodal Brain Image Analysis. Vol 8159. Lecture Notes in Computer Science. Springer International Publishing; 2013:119-128. doi:10.1007/978-3-319-02126-3_12

26. Po-Yu Kao. pykao/CT2MNI152: First release of the CT to MNI 152 space registration tool. Published online December 12, 2019. doi:10.5281/ZENODO.3572912

27. Yeh FC. Population-based tract-to-region connectome of the human brain and its hierarchical topology. Nat Commun. 2022;13(1):4933. doi:10.1038/s41467-022-32595-4

28. Van Essen DC, Ugurbil K, Auerbach E, et al. The Human Connectome Project: a data acquisition perspective. NeuroImage. 2012;62(4):2222–2231. doi:10.1016/j.neuroimage.2012.02.018

29. Salimi-Khorshidi G, Douaud G, Beckmann CF, Glasser MF, Griffanti L, Smith SM. Automatic denoising of functional MRI data: combining independent component analysis and hierarchical fusion of classifiers. NeuroImage. 2014;90:449–468. doi:10.1016/j.neuroimage.2013.11.046

30. Jimenez-Marin A, De Bruyn N, Gooijers J, et al. Multimodal and multidomain lesion network mapping enhances prediction of sensorimotor behavior in stroke patients. Sci Rep. 2022;12(1):22400. doi:10.1038/s41598-022-26945-x

31. Lundberg SM, Lee SI. A unified approach to interpreting model predictions. In: Proceedings of the 31st International Conference on Neural Information Processing Systems. NIPS’17. Curran Associates Inc.; 2017:4768–4777.

32. DeLong MR, Wichmann T. Circuits and circuit disorders of the basal ganglia. Arch Neurol. 2007;64(1):20–24. doi:10.1001/archneur.64.1.20

33. Hening W, Harrington DL, Poizner H. Basal Ganglia: Motor Functions of. In: Binder MD, Hirokawa N, Windhorst U, eds. Encyclopedia of Neuroscience. Springer Berlin Heidelberg; 2008:346–350. doi:10.1007/978-3-540-29678-2_561

34. Vicente AF, Bermudez MA, Romero MDC, Perez R, Gonzalez F. Putamen neurons process both sensory and motor information during a complex task. Brain Res. 2012;1466:70–81. doi:10.1016/j.brainres.2012.05.037

35. Erzurumlu R, Sengul G, Ulupinar E. Human Neuroanatomy. Academic Press; 2024.

36. Gómez-Ocádiz R, Silberberg G. Corticostriatal pathways for bilateral sensorimotor functions. Curr Opin Neurobiol. 2023;83:102781. doi:10.1016/j.conb.2023.102781

37. Rousseau PN, Bazin PL, Steele CJ. Multiscale gradients of corticopontine structural connectivity. Sci Rep. 2025;15(1):16399. doi:10.1038/s41598-025-00886-7

38. Terao S, Miura N, Takeda A, Takahashi A, Mitsuma T, Sobue G. Course and distribution of facial corticobulbar tract fibres in the lower brain stem. J Neurol Neurosurg Psychiatry. 2000;69(2):262–265. doi:10.1136/jnnp.69.2.262

39. Niu J, Ding L, Li JJ, et al. Modality-Based Organization of Ascending Somatosensory Axons in the Direct Dorsal Column Pathway. J Neurosci. 2013;33(45):17691–17709. doi:10.1523/JNEUROSCI.3429-13.2013

40. Bajada CJ, Lambon Ralph MA, Cloutman LL. Transport for language south of the Sylvian fissure: The routes and history of the main tracts and stations in the ventral language network. Cortex J Devoted Study Nerv Syst Behav. 2015;69:141–151. doi:10.1016/j.cortex.2015.05.011

41. Corbetta M, Patel G, Shulman GL. The reorienting system of the human brain: from environment to theory of mind. Neuron. 2008;58(3):306–324. doi:10.1016/j.neuron.2008.04.017

42. Uddin LQ, Kelly AM, Biswal BB, Castellanos FX, Milham MP. Functional connectivity of default mode network components: correlation, anticorrelation, and causality. Hum Brain Mapp. 2009;30(2):625–637. doi:10.1002/hbm.20531

43. Hordacre B, Lotze M, Jenkinson M, et al. Fronto-parietal involvement in chronic stroke motor performance when corticospinal tract integrity is compromised. NeuroImage Clin. 2021;29:102558. doi:10.1016/j.nicl.2021.102558

44. Diez I, Drijkoningen D, Stramaglia S, et al. Enhanced prefrontal functional–structural networks to support postural control deficits after traumatic brain injury in a pediatric population. Netw Neurosci. 2017;1(2):116–142. doi:10.1162/NETN_a_00007

